# Benchmarking Human-AI Collaboration for Common Evidence Appraisal Tools

**DOI:** 10.1101/2024.04.21.24306137

**Authors:** Tim Woelfle, Julian Hirt, Perrine Janiaud, Ludwig Kappos, John P. A. Ioannidis, Lars G. Hemkens

## Abstract

**Background:** It is unknown whether large language models (LLMs) may facilitate time- and resource-intensive text-related processes in evidence appraisal.

**Objectives:** To quantify the agreement of LLMs with human consensus in appraisal of scientific reporting (PRISMA) and methodological rigor (AMSTAR) of systematic reviews and design of clinical trials (PRECIS-2). To identify areas, where human-AI collaboration would outperform the traditional consensus process of human raters in efficiency.

**Design:** Five LLMs (Claude-3-Opus, Claude-2, GPT-4, GPT-3.5, Mixtral-8x22B) assessed 112 systematic reviews applying the PRISMA and AMSTAR criteria, and 56 randomized controlled trials applying PRECIS-2. We quantified agreement between human consensus and (1) individual human raters; (2) individual LLMs; (3) combined LLMs approach; (4) human-AI collaboration. Ratings were marked as deferred (undecided) in case of inconsistency between combined LLMs or between the human rater and the LLM.

**Results:** Individual human rater accuracy was 89% for PRISMA and AMSTAR, and 75% for PRECIS-2. Individual LLM accuracy was ranging from 63% (GPT-3.5) to 70% (Claude-3-Opus) for PRISMA, 53% (GPT-3.5) to 74% (Claude-3-Opus) for AMSTAR, and 38% (GPT-4) to 55% (GPT-3.5) for PRECIS-2. Combined LLM ratings led to accuracies of 75-88% for PRISMA (4-74% deferred), 74-89% for AMSTAR (6-84% deferred), and 64-79% for PRECIS-2 (18-88% deferred). Human-AI collaboration resulted in the best accuracies from 89-96% for PRISMA (25/35% deferred), 91-95% for AMSTAR (27/30% deferred), and 80-86% for PRECIS-2 (76/71% deferred).

**Conclusions:** Current LLMs alone appraised evidence worse than humans. Human-AI collaboration may reduce workload for the second human rater for the assessment of reporting (PRISMA) and methodological rigor (AMSTAR) but not for complex tasks such as PRECIS-2.

## Introduction

The assessment of reporting, methodological rigor, and design features of biomedical research is essential for evidence-based medicine. However, these evaluations require extensive resources.

Many evidence appraisal tools are primarily text-based and follow instructions of varying complexity, e.g., reporting checklists or clinical trial tools such as “Preferred Reporting Items for Systematic reviews and Meta-Analyses” (PRISMA),^1^ “A MeaSurement Tool to Assess systematic Reviews” (AMSTAR),^2^ and “PRagmatic Explanatory Continuum Indicator Summary 2” (PRECIS-2).^3^

Traditional machine learning and natural language processing methods have been explored for years to extract for example “population, intervention, control, outcome” (PICO) information or key elements used for “risk of bias” (RoB) assessments from study reports.^4,5^ More advanced deep learning applications have been applied for automatically assessing reporting on a large scale.^6^

Large language models (LLMs) such as OpenAI’s ChatGPT have gained great attention due to their advanced language-processing capabilities and presumed reasoning, beating several artificial intelligence (AI) benchmarks.^7–9^ While their potential to facilitate systematic reviews is generally acknowledged, there is an increasing discussion of their limitations and of the need for caution in using these tools.^10^ The efficacy of LLMs for screening and data extraction for systematic reviews has been highly mixed, but there appear to be scenarios where they can help with these tasks.^11,12^

We quantified the agreement of five individual LLMs with human consensus in the assessment of evidence appraisal tools of different levels of complexity: reporting (PRISMA) and methodological rigor (AMSTAR) of systematic reviews, and degree of pragmatism of clinical trials (PRECIS-2). We assessed how much complexity of assessment can be handled by current models, which one performs best, and whether the combination of multiple LLMs increases accuracy. Finally, we combined individual human raters with LLMs and evaluated whether such human-AI collaboration outperforms a traditional consensus process of multiple human raters in efficiency.

## Methods

### Selecting datasets and evidence appraisal tools

We selected datasets for which independent ratings from two human raters and their consensus were available. The independent assessment by at least two human raters is the standard in systematic reviews (e.g., for Cochrane reviews^13^), and is also commonly used to assess reporting and methodological rigor (e.g., using PRISMA^14^ and AMSTAR^14,15^), or study designs (e.g., using PRECIS-2^16^).

For reporting and methodological rigor, we used human assessments of PRISMA and AMSTAR for 112 systematic reviews and meta-analyses in the field of pediatric surgery (data kindly shared by Cullis and colleagues).^14^ PRISMA contains 27 items and AMSTAR 11 items (Supplement), rated as no / yes / not applicable (NA).^1,2^ The two raters were content experts (British pediatric surgeons).

For pragmatism in clinical trial design, we used human ratings of PRECIS-2 for 56 randomized controlled trials (RCTs) within the PragMeta database.^16,17^ PRECIS-2 contains 9 domains (Supplement), rated ordinally from 1 (very explanatory) to 5 (very pragmatic) or NA.^3^ Human rater 1 was an experienced systematic-reviewer and meta-researcher. Human rater 2 was either one of two post-graduate MSc students in epidemiology with special training in PRECIS-2 assessment or a senior clinical epidemiologist and expert in pragmatic trial design.

### Large language models

We used four proprietary LLMs (Anthropic’s Claude-3-Opus and Claude-2^18,19^ and OpenAI’s GPT-4 and GPT-3.5^20,21^) and one open source LLM (Mistral AI’s Mixtral-8x22B^22^), Table 1. As of April 2024, Anthropic’s largest model Claude-3-Opus is among the best-performing LLMs according to the popular Chatbot Arena,^23^ and available via chat for Claude Pro users. Claude-2 is an older and cheaper version. GPT-4 is OpenAI’s largest model, among the best-performing LLMs since its release in March 2023, and available via chat for ChatGPT Plus users. The older and cheaper GPT-3.5 is available as ChatGPT for free. Mistral AI’s Mixtral-8x22B is fully open source, meaning the model can be downloaded and run locally, provided the right hardware is available.

**Table 1:**
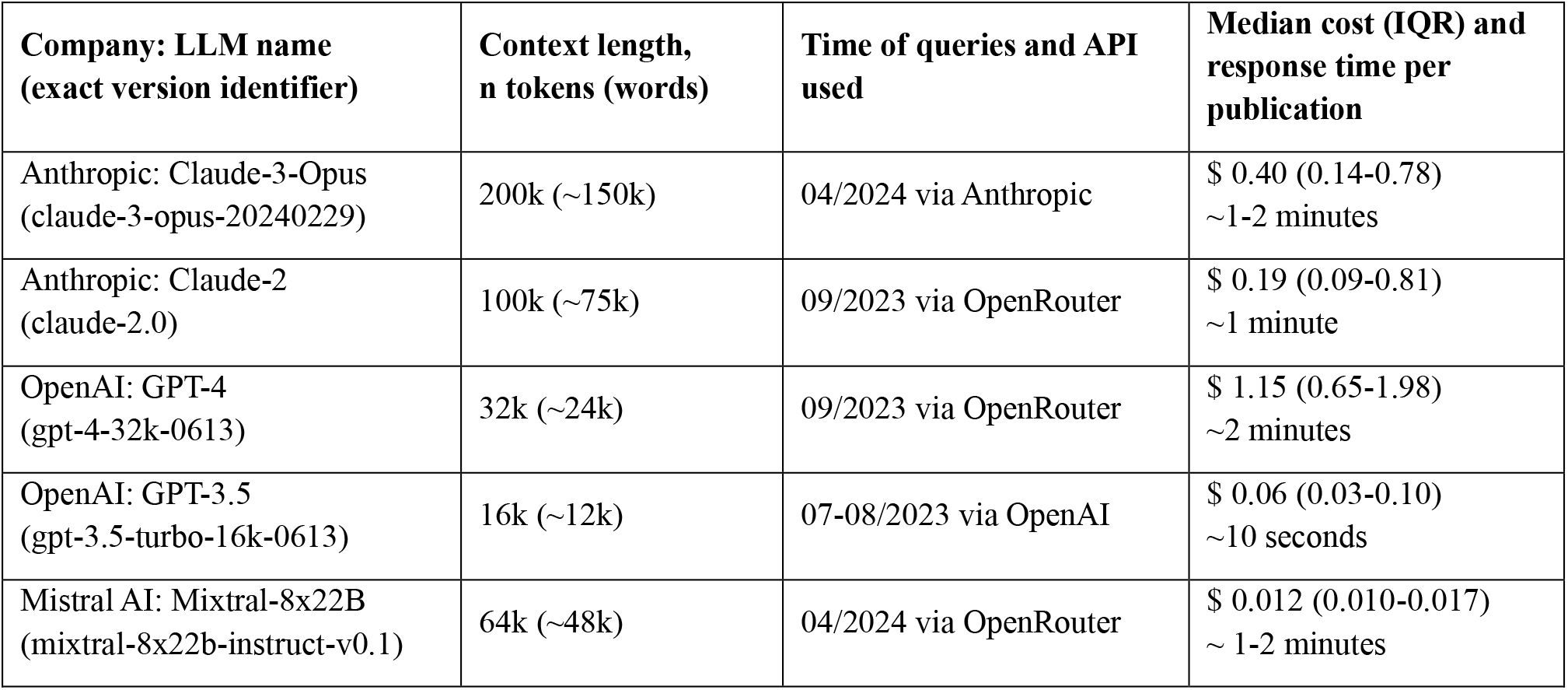
LLMs used: name, context length, timeframe of queries and API used, median cost and response time per publication. LLM: large language model. API: application programming interface.

| **Company: LLM name**<br>**(exact version identifier)** | **Context length,**<br>**n tokens (words)** | **Time of queries and API**<br>**used** | **Median cost (IQR) and**<br>**response time per**<br>**publication** |
| --- | --- | --- | --- |
| Anthropic: Claude-3-Opus<br>(claude-3-opus-20240229) | 200k (~150k) | 04/2024 via Anthropic | \$ 0.40 (0.14-0.78)<br>~1-2 minutes |
| Anthropic: Claude-2<br>(claude-2.0) | 100k (~75k) | 09/2023 via OpenRouter | \$ 0.19 (0.09-0.81)<br>~1 minute |
| OpenAI: GPT-4<br>(gpt-4-32k-0613) | 32k (~24k) | 09/2023 via OpenRouter | \$ 1.15 (0.65-1.98)<br>~2 minutes |
| OpenAI: GPT-3.5<br>(gpt-3.5-turbo-16k-0613) | 16k (~12k) | 07-08/2023 via OpenAI | \$ 0.06 (0.03-0.10)<br>~10 seconds |
| Mistral AI: Mixtral-8x22B<br>(mixtral-8x22b-instruct-v0.1) | 64k (~48k) | 04/2024 via OpenRouter | \$ 0.012 (0.010-0.017)<br>~ 1-2 minutes |

Claude-3-Opus was the only multimodal model we employed, meaning we uploaded PDF files converted to plain images and the model performed optical character recognition itself. The four other models could only process text and no images. While later versions of GPT-4 were also multimodal, the version we used (gpt-4-32k-0613) had no vision capabilities yet.

To increase comparability, reproducibility, and reliability, all models were queried via pay-per-use application programming interfaces (API), allowing us to query specific timestamped versions of each model and to set the probabilistic parameter “temperature” to zero. As stated by OpenAI: “Models are non-deterministic, meaning that identical inputs can yield different outputs. Setting temperature to 0 will make the outputs mostly deterministic, but a small amount of variability may remain”.^21^ Claude-3-Opus was directly accessed via Anthropic’s API and GPT-3.5 directly via OpenAI’s API, while Claude-2, GPT-4, and Mixtral-8x22B were accessed via the intermediary API “OpenRouter”, Table 1.^24^

### Prompt engineering

We provided each model an introduction and briefing in the “system prompt” and the publication to assess and stepwise instructions in the “user prompt” to clearly separate them. Only Mixtral-8x22B does not provide this separation, in which case we simple concatenated the two parts.

We used in-context expert impersonation to open our system prompts, which is a common prompt-engineering strategy and improves LLM performance:^25^ “You are an expert in systematic reviews” for PRISMA and AMSTAR and “[…] in clinical trial design” for PRECIS-2. After briefly explaining the task in one sentence, the following “briefing information” was provided: for PRISMA and AMSTAR, the original item-wise instructions provided by Cullis and colleagues in their supplementary S1 Dataset were used (roughly one page of text each).^14^ For PRECIS-2, Claude-3-Opus, GPT-3.5, and Mixtral-8x22B were provided with page 4 of the official toolkit, detailing the 9 domains.^26^ For exploratory reasons, Claude-2 was provided with the full text of the toolkit (8 pages),^26^ and additionally the full text of the introductory publication of PRECIS-2 (Loudon 2015, 11 pages).^3^ GPT-4 was provided with page 4 of the toolkit and the section “The domains in detail” of Loudon 2015 (pages 5-10 without boxes).^3^

The user prompts first contained one of the publications to assess (112 study reports for PRISMA and AMSTAR and 56 trial publications for PRECIS-2). For Claude-3-Opus, PDF files were exported as one PNG file per page at 150 dots per inch (DPI) and attached. For Claude-2, GPT-4, and Mixtral-8x22B, the full text was derived from the PDF top-to-bottom and attached. For GPT-3.5, because of its short context length, the full text was copied manually from PubMed Central when available and alternatively from the journal’s website: For PRISMA and AMSTAR, everything from the title to the end of the “Declaration of Interests” or “Acknowledgments” section, excluding references. Table contents were present in 63 / 112 publications, depending on the formatting of the website. For PRECIS-2, everything from the abstract to the end of the “Discussion” or “Conclusions” section was copied. Table contents were present in 43 / 56 publications. Figure and table captions were always included.

In the final paragraph of the user prompts, we employed the chain-of-thought (CoT) prompting-technique by instructing the models to perform 3 consecutive steps for each item:^27^ (1) extract relevant quotes from the publication full text, (2) explain the reasoning, and (3) give a rating in squared brackets (i.e. [Yes/No/NA] for PRISMA and AMSTAR and [Score: 1-5/NA] for PRECIS-2). All 3 consecutive steps were instructed within the same single prompt, and we never responded to the LLM’s initial response.

The CoT prompting-technique – instructing LLMs to perform intermediate steps before producing a final answer – has been shown to generally improve performance and has also been used successfully in biomedical contexts.^28^ Furthermore, it increases transparency and model interpretability. The extraction of relevant quotes is supposed to ground the models’ answers in facts and helps human raters to verify them.^29^ In fact, the extraction of relevant quotes may be a valuable feature on its own, potentially providing assistance for human raters in their assessments.

Assessment of PRISMA and AMSTAR was combined in a single prompt per publication with Claude-3-Opus, Claude-2, GPT-4, and Mixtral-8x22B and in two independent prompts per publication with GPT-3.5 due to its smaller context length. PRECIS-2 scoring was always performed in a single prompt per publication.

### Extraction of ratings and quotes

Every LLM response was saved, and ratings were extracted automatically using regular expressions. Minor formatting issues (e.g. forgotten squared brackets around ratings) or non-compliance with instructions (e.g. using unsolicited responses like “[Partial]”) were fixed automatically during extraction when possible. Major non-compliance with instructions (e.g. missing ratings) led to a minimum of 3 manual repetitions of the prompt. Both minor and major issues were quantified.

As LLMs are known to occasionally “hallucinate” false information,^29,30^ we quantified quote accuracy. Quotes within the LLM responses were extracted automatically and compared against both the full text of the publications (which the models were explicitly asked to quote) and the briefing text (which the models were not explicitly asked to quote), finding the best matching passages using the striped Smith-Waterman algorithm.^31^ For each quote and its best match, a normalized Levenshtein similarity was calculated, normalizing the minimum number of insertions and deletions necessary to transform the quote to its best match and ranging from 0 to 100%.^32^ We penalized insertions (possible “hallucinations”) twice as much as deletions (e.g. omissions of brackets).

### Analyses and outcomes

#### Agreement with human consensus

Our main outcome was agreement with human consensus measured by accuracy (agreement fraction, i.e., the proportion of identical ratings between rater and human consensus) and Cohen’s kappa. For the ordinal PRECIS-2 ratings, responses 1 and 2 (“very” and “mostly explanatory”) were pooled to “1/2” and responses 4 and 5 (“very” and “mostly pragmatic”) to “4/5” and a weighted version of Cohen’s kappa was used (Supplementary Table 1). Bootstrapping with 1000 resamples on the publication-level was performed to derive 95% confidence intervals (CIs).

We performed four analyses for each of the 3 evidence appraisal tools (PRISMA, AMSTAR, PRECIS-2) (Figure 1) while also quantifying resources used for LLMs (costs and time effort).

**Figure 1:**
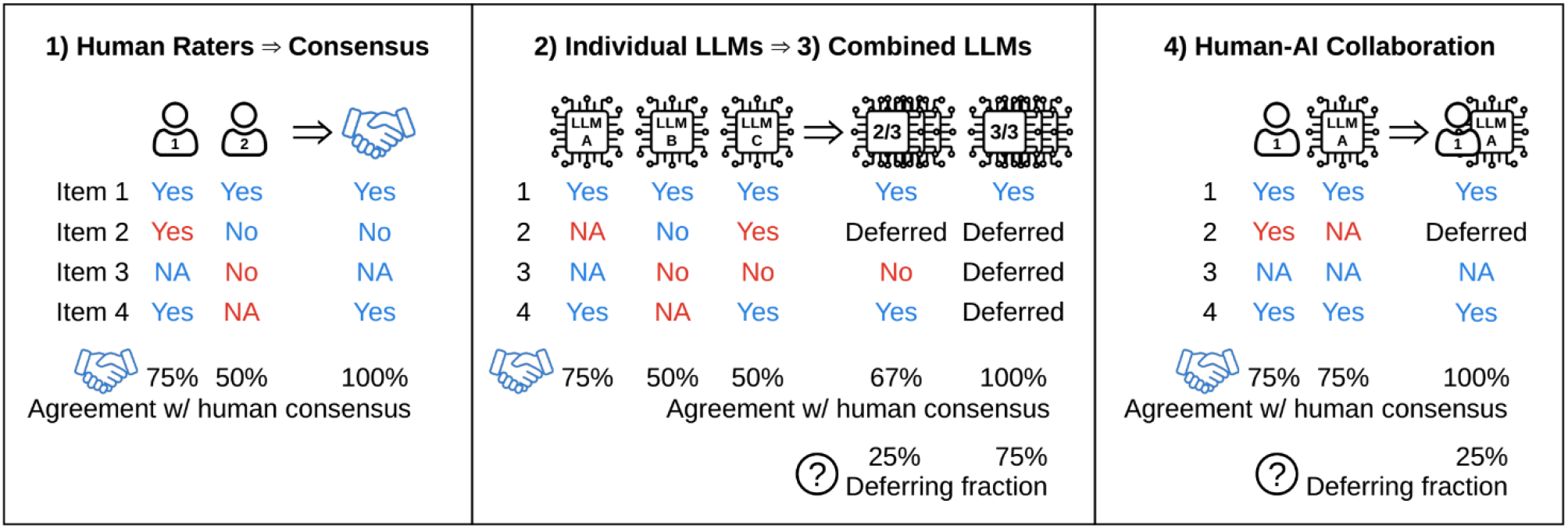
Schematic overview of the main outcome accuracy (agreement with human consensus, in blue) for a hypothetical tool with 4 items in a single publication. (1) Two human raters independently rate each item and agree on a consensus. (2) Three individual LLMs independently rate each item and are compared to human consensus. (3) They are combined using a consistency approach, using either all responses consistent in at least 2 out of 3 LLMs or all 3/3 LLMs. The proportion of inconsistent ratings is the deferring fraction and agreement is only calculated on the consistent ones. (4) A human rater is combined with an LLM: only items consistent between human and LLM are compared to human consensus and the remaining ones are deferred. LLM: large language model.

1. Human consensus vs individual human raters
2. Human consensus vs individual LLMs (Claude-3-Opus, Claude-2, GPT-4, GPT-3.5, Mixtral-8x22B)
3. Human consensus vs combined LLMs: consistency approach Combining multiple assessments from the same LLM in a “self-consistency” approach improves performance in biomedical and general contexts beyond using only a single assessment.^28,33^ We combined a total of 9 LLM assessments: 2× Claude-3-Opus, 2× Claude-2, 1× GPT-4 (due to high costs), 2× GPT-3.5, 2× Mixtral-8x22B.^34^ Deferring fraction: Inconsistent responses (i.e., without alignment among 6-12 out of the 12 LLM assessments) were considered uncertain and thus deferred to human raters, meaning the LLMs could not provide a consistent rating and a human must decide. We assumed that responses with higher consistency (i.e., among more of the 12 LLM assessments) exhibit better agreement with human consensus at the cost of higher deferring rates (as seen by others, e.g., Figure 3 in ^28^).
4. Human consensus vs human-AI collaboration We combined ratings of individual human raters with individual LLM ratings for each of the three tools. Items where the LLM aligned with the human rater were compared to human consensus. Inconsistent items were considered uncertain and thus deferred to a second human rater.

**Figure 2:**
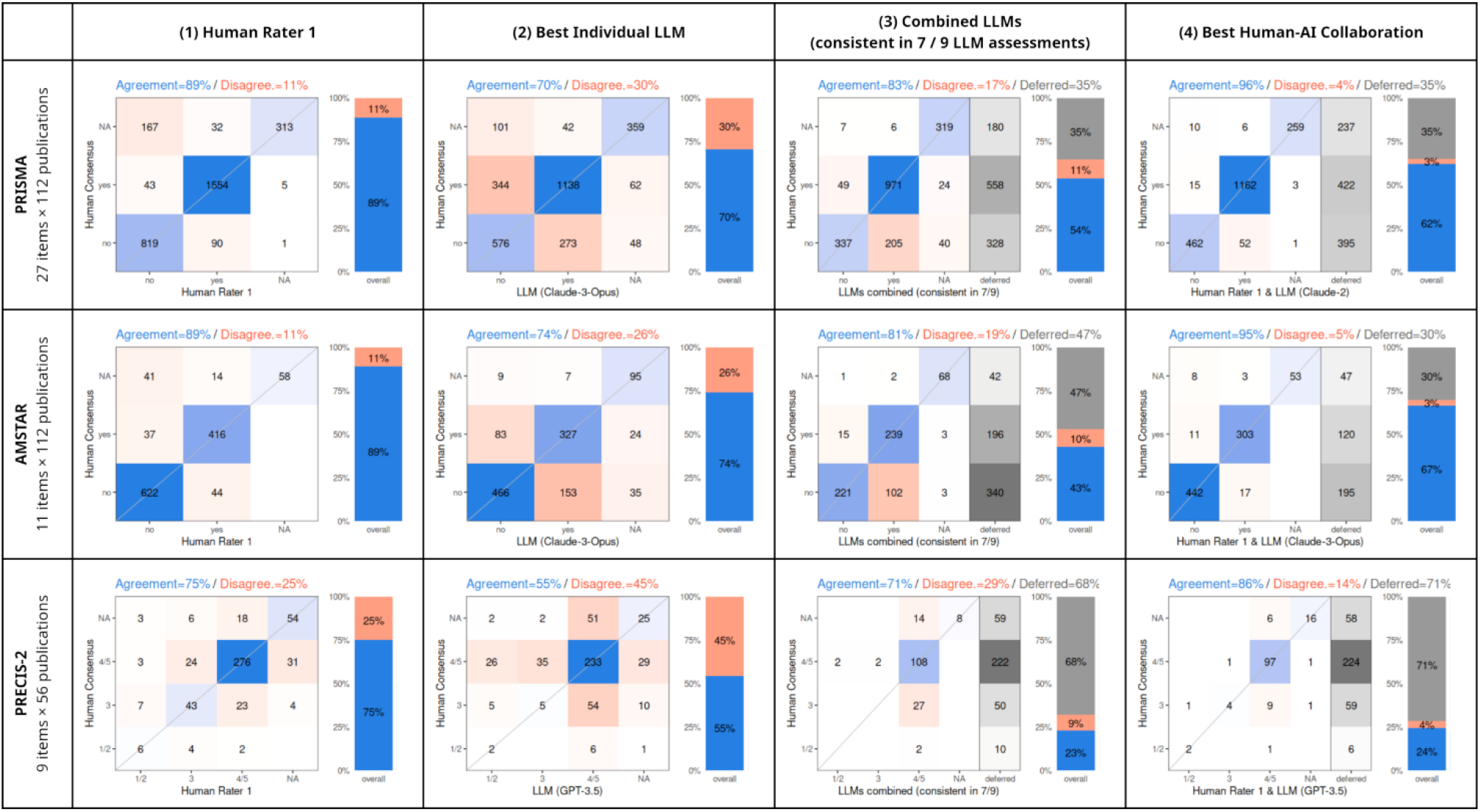
Confusion matrices for “agreement” (blue) and “disagreement” (red) with human consensus for the three tools (in rows). For 3) and 4), “deferred” (grey) refers to the proportion of responses inconsistent between LLMs or between human and LLM, which are thus considered unclear and deferred to a (second) human rater. Accuracy is only calculated for consistent responses, which thus represent a hypothetical workload reduction. LLM: large language model. PRISMA: Preferred Reporting Items for Systematic reviews and Meta-Analyses”. AMSTAR: “A MeaSurement Tool to Assess systematic Reviews”. PRECIS-2: “PRagmatic-Explanatory Continuum Indicator Summary-2”.

**Figure 3:**
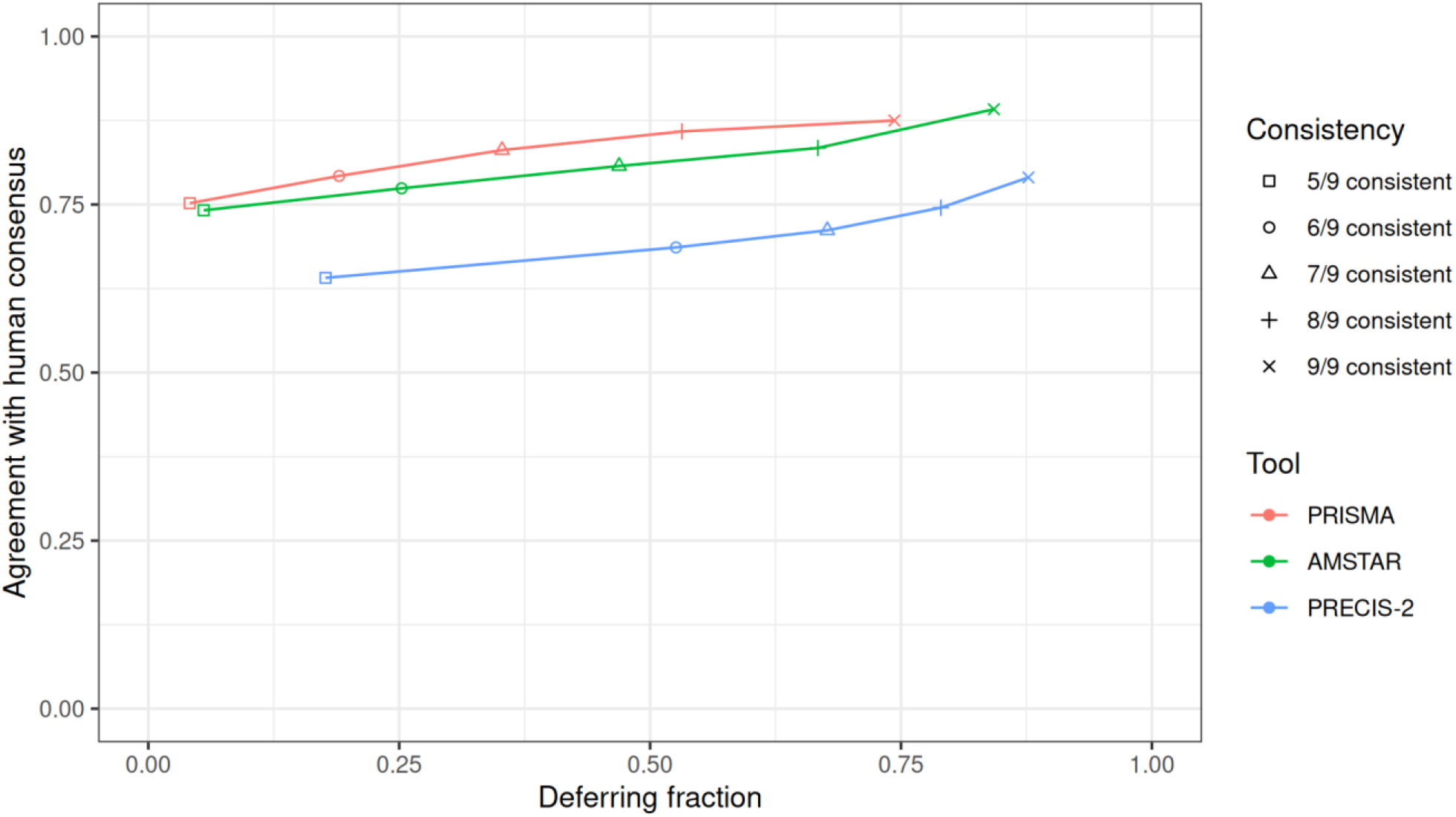
Combined LLMs through consistency approach. Combining all 9 LLM assessments per tool and using only the responses that are consistent among 5-9 out of 9 assessments led to increasing performances for all 3 tools measured by accuracy. This comes at the cost of an increasing “deferring fraction”, comprising the inconsistent responses. Combining LLMs thus allows an estimation of uncertainty.

For both (3) and (4), accuracy and kappa have to be interpreted in combination with the deferring fraction.

#### Reliability

We performed LLM prompts twice (i.e., in duplicate for Claude-3-Opus, Claude-2, GPT-4 [only 25% of publications, due to high cost], GPT-3.5, Mixtral-8x22B) and compared the ratings of each of the two runs. Due to the nature of LLMs, these duplicate runs are not independent, which is why we consider their agreement “intra-rater reliability”. All API queries were performed with minimal randomness (“temperature” 0) to allow the highest possible intra-rater reliability. We compared LLM intra-rater reliability with human inter-rater reliability.

#### Code and data availability

API querying, extraction of ratings, fixing minor formatting issues, and quantification of quote accuracy were performed in Python 3.11.4 using the parasail and rapidfuzz libraries.^32,35^ Statistical analyses and visualizations were performed in R 4.3.

Codes and data are openly available on GitHub.^36^ All prompt templates are in the Supplement. We provide a web dashboard which allows interactive exploration of results.^34^

## Results

### Agreement with human consensus

#### Individual human raters

Accuracies of human raters 1 and 2 were 89% and 90% for PRISMA, 89% and 89% for AMSTAR, and 75% and 73% for PRECIS-2, respectively (Table 2, Figure 2).

**Table 2:** Comparison of agreement between human consensus and (1) individual human rater 1; and (2) individual LLMs (Claude-3-Opus, Claude-2, GPT-4, GPT-3.5, Mixtral-8x22B) and their reliability.

| Rater | Agreement with Human Consensus |  | Reliability |  |
| --- | --- | --- | --- | --- |
|  | Cohen's kappa (95% CI) | Agreement (%<br>95% CI) | Cohen's kappa (95% CI) | Agreement (%<br>95% CI) |
| **PRISMA (27 items × 112 publications = up to 3024 ratings)** |  |  |  |  |
| (1) Human Rater 1 | 0.81 (0.78-0.83) | 2686 / 3024 (89%, 87-90%) | 0.84 (0.80-0.88)<sup>a</sup> | 2384 / 2619 (91%, 89-93%)<sup>a</sup> |
| (2) Claude-3-Opus | 0.51 (0.47-0.55)<sup>b</sup> | 2073 / 2943 (70%, 68-72%)<sup>b</sup> | 0.68 (0.64-0.71)<sup>b</sup> | 2318 / 2889 (80%, 78-82%)<sup>b</sup> |
| (2) Claude-2 | 0.51 (0.47-0.55) | 2116 / 3024 (70%, 68-72%) | 0.83 (0.80-0.85) | 2695 / 3024 (89%, 87-91%) |
| (2) GPT-4 | 0.45 (0.42-0.48)<sup>b</sup> | 2018 / 2943 (69%, 66-71%)<sup>b</sup> | 0.80 (0.72-0.86)<sup>c</sup> | 677 / 756 (90%, 85-93%)<sup>c</sup> |
| (2) GPT-3.5 | 0.40 (0.37-0.44)<sup>b</sup> | 1868 / 2943 (63%, 62-66%)<sup>b</sup> | 0.90 (0.87-0.93)<sup>b</sup> | 2762 / 2943 (94%, 92-96%)<sup>b</sup> |
| (2) Mixtral-8x22B | 0.42 (0.37-0.47)<sup>b</sup> | 1922 / 2997 (64%, 61-67%)<sup>b</sup> | 0.46 (0.40-0.52)<sup>b</sup> | 1997 / 2997 (67%, 63-70%)<sup>b</sup> |
| **AMSTAR (11 items × 112 publications = up to 1232 ratings)** |  |  |  |  |
| (1) Human Rater 1 | 0.80 (0.76-0.83) | 1096 / 1232 (89%, 87-91%) | 0.77 (0.72-0.82)<sup>a</sup> | 936 / 1067 (88%, 85-90%)<sup>a</sup> |
| (2) Claude-3-Opus | 0.56 (0.52-0.60)<sup>b</sup> | 888 / 1199 (74%, 72-76%)<sup>b</sup> | 0.68 (0.63-0.72)<sup>b</sup> | 947 / 1177 (80%, 78-83%)<sup>b</sup> |
| (2) Claude-2 | 0.39 (0.34-0.43) | 777 / 1232 (63%, 60-66%) | 0.91 (0.87-0.94) | 1163 / 1232 (94%, 92-96%) |
| (2) GPT-4 | 0.45 (0.40-0.51)<sup>b</sup> | 836 / 1199 (70%, 67-73%)<sup>b</sup> | 0.79 (0.68-0.87)<sup>c</sup> | 274 / 308 (89%, 83-94%)<sup>c</sup> |
| (2) GPT-3.5 | 0.27 (0.22-0.32) | 652 / 1232 (53%, 50-56%) | 0.79 (0.73-0.83) | 1067 / 1232 (87%, 83-90%) |
| (2) Mixtral-8x22B | 0.34 (0.29-0.39)<sup>b</sup> | 726 / 1221 (59%, 56-63%)<sup>b</sup> | 0.44 (0.38-0.50)<sup>b</sup> | 784 / 1221 (64%, 60-68%)<sup>b</sup> |
| **PRECIS-2 (9 items × 56 publications = up to 504 ratings)** |  |  |  |  |
| (1) Human Rater 1 | 0.57 (0.48-0.65) | 379 / 504 (75%, 70-80%) | 0.29 (0.20-0.37) | 288 / 504 (57%, 51-63%) |
| (2) Claude-3-Opus | 0.12 (0.06-0.19) | 225 / 504 (45%, 40-49%) | 0.65 (0.57-0.71) | 364 / 504 (72%, 68-77%) |
| (2) Claude-2 | 0.01 (-0.08-0.08) | 222 / 504 (44%, 39-49%) | 0.71 (0.63-0.77) | 384 / 504 (76%, 71-81%) |
| (2) GPT-4 | 0.02 (-0.04-0.08) | 193 / 504 (38%, 33-43%) | 0.68 (0.52-0.80)<sup>c</sup> | 80 / 126 (63%, 52-75%)<sup>c</sup> |
| (2) GPT-3.5 | 0.04 (-0.05-0.12)<sup>b</sup> | 265 / 486 (55%, 50-58%)<sup>b</sup> | 0.62 (0.52-0.71)<sup>b</sup> | 370 / 477 (78%, 71-84%)<sup>b</sup> |
| (2) Mixtral-8x22B | 0.05 (-0.03-0.13) | 244 / 504 (48%, 43-54%) | 0.40 (0.29-0.50) | 305 / 504 (61%, 54-67%) |

a: for human rater 2 ratings for 15 publications missing for PRISMA and AMSTAR
b: responses for 1-3 publications unsuccessful and thus missing
c: GPT-4 intra-rater reliability based on 25% of publications (28 for PRISMA & AMSTAR, 14 for PRECIS-2)

#### Individual LLMs

For PRISMA, individual LLM accuracies ranged from 63% (GPT-3.5) to 70% (Claude-3-Opus), for AMSTAR from 53% (GPT-3.5) to 74% (Claude-3-Opus), and for PRECIS-2 from 38% (GPT-4) to 55% (GPT-3.5) (Table 2, Figure 2). The Supplementary Prompt Templates contain an exemplary GPT-4 prompt and response for PRECIS-2.

While differences in the prompts used complicate comparisons between individual LLMs, averaging their performance nevertheless gives a broad overview: The best average accuracy was achieved by Claude-3-Opus (63%), followed by Claude-2 (59%), GPT-4 (59%), Mixtral-8x22B (57%), and GPT-3.5 (57%). This ranking is also reflected by their average kappas: 0.40, 0.30, 0.31, 0.27, and 0.24, respectively.

#### Combined LLMs: Consistency approach

All 9 LLM assessments were combined using only ratings consistent in a majority of LLM assessments. Responses without such a consistent majority would be deferred to human raters. This approach led to substantial improvements with accuracies ranging from 75-88% for PRISMA (while deferring 4-74% of ratings), from 74-89% for AMSTAR (while deferring 6-84% of ratings), and from 64-79% for PRECIS-2 (with deferring fractions from 18-88%) (Figure 2). By consecutively increasing the consistency threshold from ≥5/9 to finally 9/9, accuracies and deferring fractions increased gradually (Figure 3). The performance of combined LLMs at high consistency-thresholds (9/9 for PRISMA and AMSTAR and ≥8/9 for PRECIS-2) was statistically indifferent from human performance, evidenced by overlapping confidence intervals for both accuracy and kappa. (Table 3).

**Table 3:**
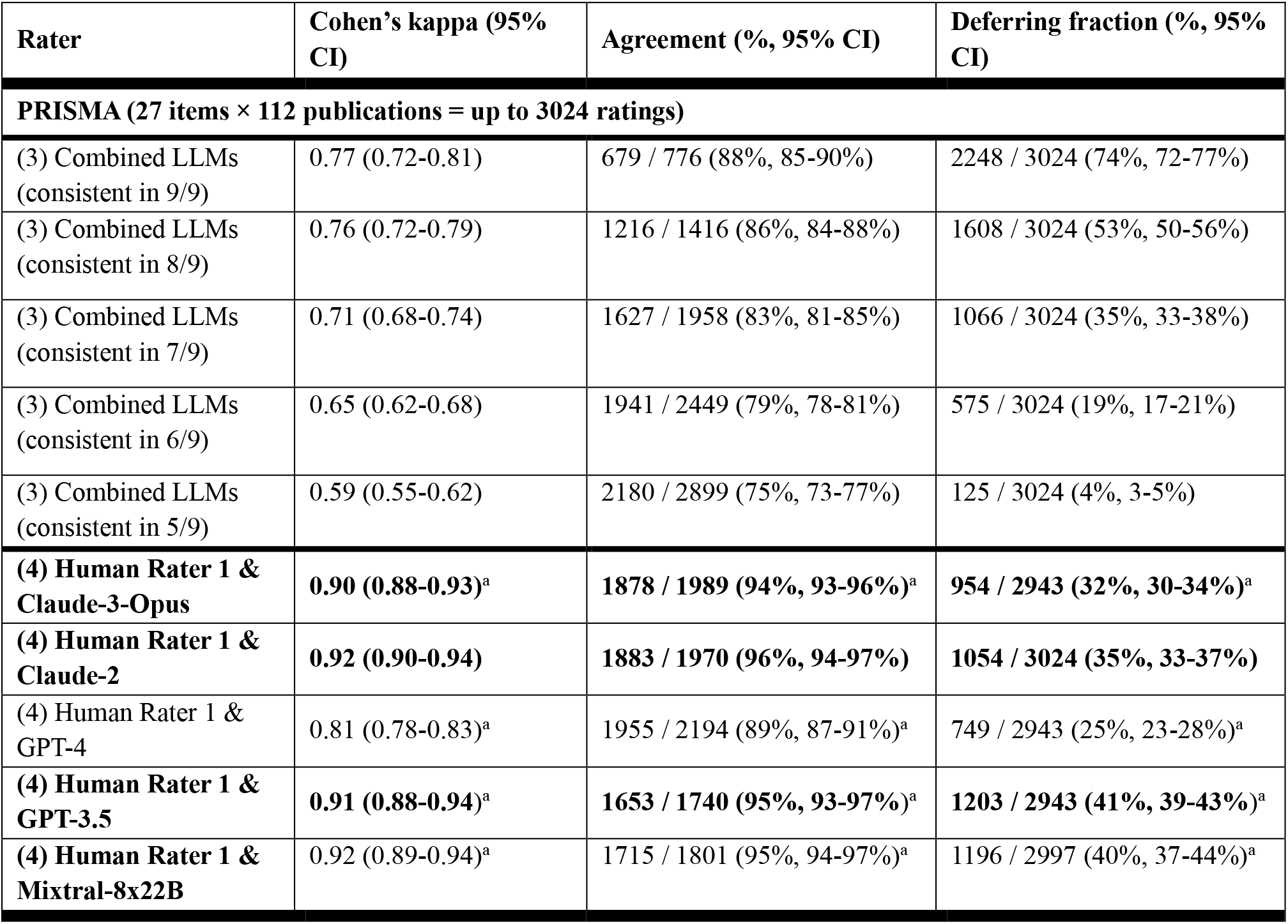
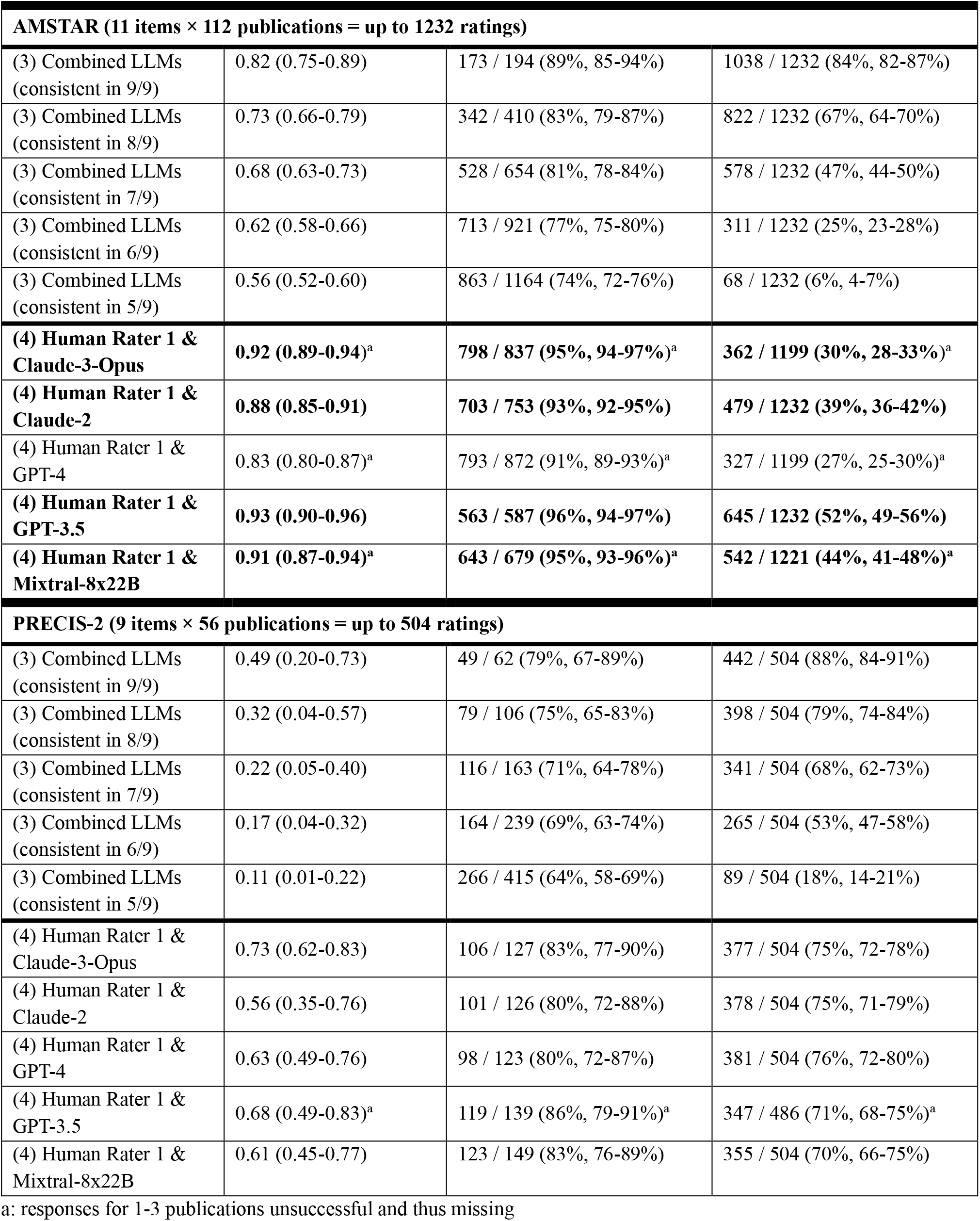
Comparison of agreement between human consensus and (3) combined LLMs (consistent in 5-9 of 9 LLM assessments), (4) human-AI collaboration for each of the three tools. Bold cells are significantly better than the human rater alone (compare Table 1).

| Rater | Cohen's kappa (95% CI) | Agreement (%, 95% CI) | Deferring fraction (%, 95% CI) |
| --- | --- | --- | --- |
| **PRISMA (27 items × 112 publications = up to 3024 ratings)** |  |  |  |
| (3) Combined LLMs (consistent in 9/9) | 0.77 (0.72-0.81) | 679 / 776 (88%, 85-90%) | 2248 / 3024 (74%, 72-77%) |
| (3) Combined LLMs (consistent in 8/9) | 0.76 (0.72-0.79) | 1216 / 1416 (86%, 84-88%) | 1608 / 3024 (53%, 50-56%) |
| (3) Combined LLMs (consistent in 7/9) | 0.71 (0.68-0.74) | 1627 / 1958 (83%, 81-85%) | 1066 / 3024 (35%, 33-38%) |
| (3) Combined LLMs (consistent in 6/9) | 0.65 (0.62-0.68) | 1941 / 2449 (79%, 78-81%) | 575 / 3024 (19%, 17-21%) |
| (3) Combined LLMs (consistent in 5/9) | 0.59 (0.55-0.62) | 2180 / 2899 (75%, 73-77%) | 125 / 3024 (4%, 3-5%) |
| **(4) Human Rater 1 & Claude-3-Opus** | **0.90 (0.88-0.93)<sup>a</sup>** | **1878 / 1989 (94%, 93-96%)<sup>a</sup>** | **954 / 2943 (32%, 30-34%)<sup>a</sup>** |
| **(4) Human Rater 1 & Claude-2** | **0.92 (0.90-0.94)** | **1883 / 1970 (96%, 94-97%)** | **1054 / 3024 (35%, 33-37%)** |
| (4) Human Rater 1 & GPT-4 | 0.81 (0.78-0.83)<sup>a</sup> | 1955 / 2194 (89%, 87-91%)<sup>a</sup> | 749 / 2943 (25%, 23-28%)<sup>a</sup> |
| **(4) Human Rater 1 & GPT-3.5** | **0.91 (0.88-0.94)<sup>a</sup>** | **1653 / 1740 (95%, 93-97%)<sup>a</sup>** | **1203 / 2943 (41%, 39-43%)<sup>a</sup>** |
| **(4) Human Rater 1 & Mixtral-8x22B** | **0.92 (0.89-0.94)<sup>a</sup>** | **1715 / 1801 (95%, 94-97%)<sup>a</sup>** | **1196 / 2997 (40%, 37-44%)<sup>a</sup>** |

| **AMSTAR (11 items × 112 publications = up to 1232 ratings)** |  |  |  |
| --- | --- | --- | --- |
| (3) Combined LLMs<br>(consistent in 9/9) | 0.82 (0.75-0.89) | 173 / 194 (89%, 85-94%) | 1038 / 1232 (84%, 82-87%) |
| (3) Combined LLMs<br>(consistent in 8/9) | 0.73 (0.66-0.79) | 342 / 410 (83%, 79-87%) | 822 / 1232 (67%, 64-70%) |
| (3) Combined LLMs<br>(consistent in 7/9) | 0.68 (0.63-0.73) | 528 / 654 (81%, 78-84%) | 578 / 1232 (47%, 44-50%) |
| (3) Combined LLMs<br>(consistent in 6/9) | 0.62 (0.58-0.66) | 713 / 921 (77%, 75-80%) | 311 / 1232 (25%, 23-28%) |
| (3) Combined LLMs<br>(consistent in 5/9) | 0.56 (0.52-0.60) | 863 / 1164 (74%, 72-76%) | 68 / 1232 (6%, 4-7%) |
| **(4) Human Rater 1 &**<br>**Claude-3-Opus** | **0.92 (0.89-0.94)<sup>a</sup>** | **798 / 837 (95%, 94-97%)<sup>a</sup>** | **362 / 1199 (30%, 28-33%)<sup>a</sup>** |
| **(4) Human Rater 1 &**<br>**Claude-2** | **0.88 (0.85-0.91)** | **703 / 753 (93%, 92-95%)** | **479 / 1232 (39%, 36-42%)** |
| (4) Human Rater 1 &<br>GPT-4 | 0.83 (0.80-0.87)<sup>a</sup> | 793 / 872 (91%, 89-93%)<sup>a</sup> | 327 / 1199 (27%, 25-30%)<sup>a</sup> |
| **(4) Human Rater 1 &**<br>**GPT-3.5** | **0.93 (0.90-0.96)** | **563 / 587 (96%, 94-97%)** | **645 / 1232 (52%, 49-56%)** |
| **(4) Human Rater 1 &**<br>**Mixtral-8x22B** | **0.91 (0.87-0.94)<sup>a</sup>** | **643 / 679 (95%, 93-96%)<sup>a</sup>** | **542 / 1221 (44%, 41-48%)<sup>a</sup>** |
| **PRECIS-2 (9 items × 56 publications = up to 504 ratings)** |  |  |  |
| (3) Combined LLMs<br>(consistent in 9/9) | 0.49 (0.20-0.73) | 49 / 62 (79%, 67-89%) | 442 / 504 (88%, 84-91%) |
| (3) Combined LLMs<br>(consistent in 8/9) | 0.32 (0.04-0.57) | 79 / 106 (75%, 65-83%) | 398 / 504 (79%, 74-84%) |
| (3) Combined LLMs<br>(consistent in 7/9) | 0.22 (0.05-0.40) | 116 / 163 (71%, 64-78%) | 341 / 504 (68%, 62-73%) |
| (3) Combined LLMs<br>(consistent in 6/9) | 0.17 (0.04-0.32) | 164 / 239 (69%, 63-74%) | 265 / 504 (53%, 47-58%) |
| (3) Combined LLMs<br>(consistent in 5/9) | 0.11 (0.01-0.22) | 266 / 415 (64%, 58-69%) | 89 / 504 (18%, 14-21%) |
| (4) Human Rater 1 &<br>Claude-3-Opus | 0.73 (0.62-0.83) | 106 / 127 (83%, 77-90%) | 377 / 504 (75%, 72-78%) |
| (4) Human Rater 1 &<br>Claude-2 | 0.56 (0.35-0.76) | 101 / 126 (80%, 72-88%) | 378 / 504 (75%, 71-79%) |
| (4) Human Rater 1 &<br>GPT-4 | 0.63 (0.49-0.76) | 98 / 123 (80%, 72-87%) | 381 / 504 (76%, 72-80%) |
| (4) Human Rater 1 &<br>GPT-3.5 | 0.68 (0.49-0.83)<sup>a</sup> | 119 / 139 (86%, 79-91%)<sup>a</sup> | 347 / 486 (71%, 68-75%)<sup>a</sup> |
| (4) Human Rater 1 &<br>Mixtral-8x22B | 0.61 (0.45-0.77) | 123 / 149 (83%, 76-89%) | 355 / 504 (70%, 66-75%) |

a: responses for 1-3 publications unsuccessful and thus missing

#### Human-AI collaboration

For PRISMA, combining human rater 1 with individual LLMs by only considering ratings consistent between human and LLM led to accuracies ranging from 89% (GPT-4) to 96% (Claude-2) while deferring 25% and 35% of responses to the second human rater, respectively. Conversely, this would spare the second human rater 65% of responses when accepting 96% accuracy, i.e., one wrong response for every ∼25 responses spared. Human-AI collaboration with Claude-3-Opus, Claude-2, GPT-3.5, and Mixtral-8x22B led to significantly more accurate responses than either human rater 1 or 2 alone (8/10 possible human-AI pairs, Table 3, Figure 2, Supplementary Table 2).

For AMSTAR, human-AI collaboration led to accuracies ranging from 91% (GPT-4) to 95% (Claude-3-Opus) while deferring 27% and 30% of responses, respectively. Conversely, this would spare the second human rater 70% responses when accepting 95% accuracy, i.e., one wrong response for every ∼20 responses spared. Human-AI collaboration with Claude-3-Opus, Claude-2, GPT-3.5, and Mixtral-8x22B led to significantly more accurate responses than either human rater 1 or 2 alone (8/10 possible human-AI pairs, Table 3, Figure 2, Supplementary Table 2).

For PRECIS-2, accuracies were ranging from 80% (GPT-4) to 86% (GPT-3.5) while deferring 76% and 71% of responses, respectively. Conversely, this would spare the second human rater 29% of responses when accepting 86% accuracy, i.e., one wrong response for every ∼7 responses spared. Only the combination of human rater 2 with GPT-3.5 led to significantly more accurate responses than human rater 2 alone (1/10 possible human-AI pairs, Table 3, Figure 2, Supplementary Table 2).

In all cases, median agreements with human consensus and deferring fractions for individual publications and for individual items were very similar to the overall metrics.

### Reliability

Human inter-rater reliability measured by agreement was 91%, 88%, 57% and by kappa 0.84, 0.77, 0.29 for PRISMA, AMSTAR, PRECIS-2, respectively.

For PRISMA, intra-rater reliability of LLMs was similar to human inter-rater-reliability for Claude-2, GPT-4, and GPT-3.5 but worse for Claude-3-Opus and Mixtral-8x22B. For AMSTAR, it was better than humans for Claude-2, similar for GPT-4 and GPT-3.5, but worse for Claude-3-Opus and Mixtral-8x22B. For PRECIS-2, it was better than humans for Claude-3-Opus, Claude-2, GPT-4, and GPT-3.5, and similar for Mixtral-8x22B (Table 2).

### Cost, effort, formatting and quoting accuracy

Mixtral-8x22B was the most affordable model with a median of $1.20 per 100 papers and GPT-4 the most expensive one with a median of $115.00. Model response speeds were ranging from ∼10 seconds (GPT-3.5) to 2 minutes (GPT-4) per paper (Table 1). Rarely, API rate limits required a break until the end of the day to continue calling Claude-3-Opus, Claude-2, and GPT-4.

Claude-3-Opus could persistently not process 3/112 (2.7%) publications for PRISMA and AMSTAR because of the output being blocked by Anthropic’s content filtering policy or being too long. Claude-2 was able to process all publications. GPT-4 could not process 3 lengthy publications (2.7%) for PRISMA and AMSTAR combined due to its context length. GPT-3.5 could persistently not process 3/112 publications (2.7%) for PRISMA and 2/56 publications (3.6%) for PRECIS-2. Mixtral-8x22B could persistently not process 1/112 publications (1%) for PRISMA and AMSTAR. Claude-3-Opus, GPT-3.5, and Mixtral-8x22B had to be re-prompted several times for up to 13% of publications until success. Claude-3-Opus and Claude-2 rarely and GPT-3.5 and Mixtral-8x22B often exhibited minor automatically fixable formatting issues like forgotten squared brackets around ratings or wrong responses like “[Unclear]” (Supplementary Table 3).

All prompts required the LLMs to “extract 1-3 relevant quotes from the full text” per item. For PRISMA (27 items), a median of 14 quotes (range 5-17) were provided per publication, 0.5 quotes/item. For AMSTAR (11 items), a median of 7 quotes (range 4-8) were provided per publication, 0.6 quotes/item. For PRECIS-2 (9 domains), a median of 10 quotes (range 9-10) were provided per publication, 1 quote/domain. Claude-3-Opus, Claude-2, and Mixtral-8x22B sometimes quoted from the provided briefings instead of the full text to be assessed, which was not part of the instructions, while GPT-4 and GPT-3.5 rarely did this. The median quote similarity with the original full text was 99%, with some quotes slightly shortened (e.g., removing references or brackets) or rephrased (Supplementary Table 3).

## Discussion

This analysis of the agreement of LLMs with human consensus in evidence appraisal of different levels of complexity identified some areas where human-AI collaboration might outperform a traditional consensus process of human raters in efficiency but also indicated clear limitations.

Individual LLMs performed significantly worse than humans for all three evidence appraisal tools. They generally performed best for reporting assessment with PRISMA, followed by the assessment of systematic review methodological rigor with AMSTAR. For the assessment of clinical trial pragmatism through PRECIS-2, the accuracy was poor and not useful in research practice. This graded performance likely reflects the increasing complexity between these tasks. Reporting assessment with PRISMA is almost exclusively a task of text and language assessment, thus an optimal scenario for LLMs. AMSTAR uses (like many risk-of-bias tools) checklists and questions where specific words and phrases are highly indicative of methodological features (e.g., “systematic search”, “grey literature”) and sufficient for a superficial categorization. Contrarily, pragmatism of clinical trials (PRECIS-2) is determined by complex features, and it requires more than a search for specific signal words and phrases. This increasing complexity may also be reflected by the decreasing human inter-rater reliability with 91% for PRISMA, 88% for AMSTAR, and 57% for PRECIS-2.

Anthropic’s newest, largest, and costliest model Claude-3-Opus performed best for PRISMA and AMSTAR, followed by Claude-2 and GPT-4, while the smallest and cheapest models GPT-3.5 and Mixtral-8x22B performed worst. Interestingly however, this ranking was somewhat inverted for PRECIS-2: Claude-3-Opus, Claude-2, and GPT-4 performed worse than GPT-3.5 and Mixtral-8x22B. We believe differences in prompts, model “personality”, and class imbalance to explain this counterintuitive paradox: The dataset used to assess PRECIS-2 contained mostly pragmatic trials and few explanatory ones. The smaller, simpler models might somehow be biased towards more pragmatic scores, compared to the otherwise better performing larger models Claude-3-Opus, Claude-2, and GPT-4, which might be trained to be more balanced. Of note, the fully open source Mixtral-8x22B on average performed better than the proprietary GPT-3.5.

Combining all LLM assessments led to increasing accuracies and deferring fractions with increasing consistency thresholds. At the highest consistency-thresholds, performance reached that of human raters, albeit only for few responses due to very high deferring fractions. Selecting only consistent majority ratings and filtering out inconsistent ones can also be regarded as a measure of uncertainty, which is very important for artificial intelligence algorithms.^37^ The increasing deferring fractions may also reflect the increasing complexity between the three tools.

Overall, these levels of accuracy suggest that individual and combined LLMs with our prompts are not yet good enough to be used alone in these evidence appraisals, and their performance is worse in more complex situations.

The best performance and currently most promising approach was human-AI collaboration: ratings consistent between a human rater and LLMs showed significantly better accuracies than human raters alone for 8/10 possible collaboration-pairs for PRISMA and AMSTAR (up to 96% accuracy for PRISMA and 95% for AMSTAR compared to 89% accuracy of human rater 1 alone). Approximately one-third of the items would still need to be assessed by a second human rater, but sparing two-thirds of the items might mean substantial workload reduction for the second human rater. Most models could thus be used to identify high-certainty ratings by the first human rater and filter out low-certainty ones to be deferred and double-checked. The second human rater would only have to rate these deferred items, leading to potential workload reduction. For PRECIS-2, there was still substantial improvement in accuracy but statistically significant in only 1/10 collaboration-pairs and a large majority of items would still need to be assessed by the second human rater.

Today’s LLMs may thus already provide a viable research partner for human raters for PRISMA and AMSTAR, but the exact reduction in workload is difficult to estimate. We cannot exclude that the reduction may not be that substantial, e.g., if the context of the non-deferred items needs to be read and understood anyway before the deferred items are also appraised.

### Limitations

First, there is a general concern about “train/test contamination” or “data leakage” with all LLM benchmarks because of the extensive web scraping used for their training. This refers to the risk of machine learning models having seen and potentially memorized test-outcomes from training-data. However, this typically affects popular data found multiple times on the internet, as it has been shown that “duplication encourages memorization”.^38,39^ While the human consensus datasets used as comparators in this work are openly available on the internet, they can only be found as tabular data (Excel/CSV). Tabular data are unlikely to be part of an LLM training corpus, as they are not useful in self-supervised training to predict the next word of a sentence. Even if they were part of an LLM training corpus, memorization would be highly unlikely because of very low numbers of duplications. Nevertheless, only prospective replication of these results with new human consensus datasets would eliminate the risk of train/test contamination.

Second, as discussed above, the dataset used in this study to assess PRECIS-2 contains mostly pragmatic trials and few explanatory ones. A more balanced dataset could complement the current findings.

Third, two of the three tools used have seen updates in recent years: PRISMA 2020^40^ and AMSTAR-2^41^, meaning the current results might only partly apply to them. However, differences between the versions that we used and the updated versions are not major.

Fourth, the presented datasets contain ratings from only two human experts, whereas a higher number would make the human consensus more robust and eventually approximate a “ground truth” – a strong term which cannot be claimed for the presented datasets. However, two human raters present the most common setting and thus reflect real research practice. Future, prospective work would benefit from having many human raters with the highest expertise to maximize their accuracy.

Finally, except Claude-3-Opus, the LLMs used could only process text and were blind to images. Figures potentially relevant for PRISMA, AMSTAR, and PRECIS-2 include flowcharts and sometimes tables are also presented as images. Newer versions of GPT-4 released after our assessment also allow multimodal prompts^20^.

### Outlook

We provide an open and reproducible benchmark that can be built upon by other groups. There are many directions that may further improve performance, e.g. testing more LLMs or more diverse prompt engineering techniques, ranging from minor variations in wording to more sophisticated multi-prompt-techniques like tree-of-thoughts and graph-of-thoughts.^42,43^ Furthermore, both industry and academia have introduced multimodal LLMs fine-tuned specifically for biomedical contexts, demonstrating encouraging performance. ^44,45^ However, these models are not generally available. Fine-tuning existing models towards a single evidence appraisal tool may also yield benefits.

Finally, the presented framework can be extended to other evidence appraisal tools like “CONsolidated Standards Of Reporting Trials” (CONSORT), for which a short report evaluating ChatGPT was published,^46^ or “Risk of Bias” (RoB), for which a protocol intending similar evaluations was published.^47^

## Conclusion

Current LLMs alone appraised evidence substantially worse than humans. Pairing a first human rater with an LLM as human-AI collaboration may reduce workload for the second human rater for the assessment of reporting (PRISMA) and methodological rigor (AMSTAR) while maintaining very high accuracy but not for more complex tasks such as assessing pragmatism of clinical trials (PRECIS-2).

## Data Availability

All codes and data are openly available on GitHub at https://github.com/timwoelfle/Evidence-Appraisal-AI, reference number 36.

https://timwoelfle.github.io/Evidence-Appraisal-AI/

## Acknowledgment

We sincerely thank Paul Stephen Cullis and colleagues for providing individual human ratings for PRISMA and AMSTAR for their systematic review in addition to the already openly shared human consensus data.^14^

## Supplementary Tables

**Supplementary Table 1:** Cohen’s kappa weight matrix for pooled PRECIS-2. Responses 1 and 2 (“very” and “mostly explanatory”) and responses 4 and 5 (“very” and “mostly pragmatic”) were pooled.

|  | 1 / 2 | 3 | 4 / 5 | NA | deferred |
| --- | --- | --- | --- | --- | --- |
| 1 / 2 | 0 | 1 | 4 | 1 | 0 |
| 3 | 1 | 0 | 1 | 1 | 0 |
| 4 / 5 | 4 | 1 | 0 | 1 | 0 |
| NA | 1 | 1 | 1 | 0 | 0 |
| deferred | 0 | 0 | 0 | 0 | 0 |

**Supplementary Table 2:** Human-AI collaboration. Agreements between human consensus and human raters combined with individual LLMs (Claude-3-Opus, Claude-2, GPT-4, GPT-3.5, Mixtral) for each of the three tools. Bold cells are significantly better than the human rater alone (italic). CI: 95% confidence interval.

|  | Human Rater 1 |  |  | Human Rater 2 |  |  |
| --- | --- | --- | --- | --- | --- | --- |
|  | Cohen's kappa (CI) | Agreement (%<br>CI) | Deferring fraction (%<br>CI) | Cohen's kappa (CI) | Agreement (%<br>CI) | Deferring fraction (%<br>CI) |
| **PRISMA (27 items × 112 publications = up to 3024 ratings)** |  |  |  |  |  |  |
| *Human Rater 1 alone* | *0.81 (0.78-0.83)* | *2686 / 3024 (89%, 87-90%)* |  | *0.83 (0.81-0.86)* | *2367 / 2619 (90%, 89-92%)* |  |
| **Claude-3-Opus** | **0.90 (0.88-0.93)** | **1878 / 1989 (94%, 93-96%)** | 954 / 2943 (32%, 30-34%) | **0.93 (0.91-0.95)** | **1664 / 1735 (96%, 95-97%)** | 830 / 2565 (32%, 30-34%) |
| **Claude-2** | **0.92 (0.90-0.94)** | **1883 / 1970 (96%, 94-97%)** | 1054 / 3024 (35%, 33-37%) | **0.95 (0.93-0.96)** | **1649 / 1702 (97%, 96-98%)** | 917 / 2619 (35%, 33-37%) |
| GPT-4 | 0.81 (0.78-0.83) | 1955 / 2194 (89%, 87-91%) | 749 / 2943 (25%, 23-28%) | 0.86 (0.83-0.88) | 1710 / 1861 (92%, 90-93%) | 677 / 2538 (27%, 25-29%) |
| **GPT-3.5** | **0.91 (0.88-0.94)** | **1653 / 1740 (95%, 93-97%)** | 1203 / 2943 (41%, 39-43%) | **0.94 (0.92-0.96)** | **1472 / 1524 (97%, 95-98%)** | 1014 / 2538 (40%, 38-42%) |
| **Mixtral-8x22B** | **0.92 (0.89-0.94)** | **1715 / 1801 (95%, 94-97%)** | 1196 / 2997 (40%, 37-44%) | **0.95 (0.93-0.97)** | **1520 / 1565 (97%, 96-98%)** | 1027 / 2592 (40%, 36-43%) |
| **AMSTAR (11 items × 112 publications = up to 1232 ratings)** |  |  |  |  |  |  |
| *Human Rater 1 alone* | *0.80 (0.76-0.83)* | *1096 / 1232 (89%, 87-91%)* |  | *0.80 (0.76-0.83)* | *949 / 1067 (89%, 87-91%)* |  |
| **Claude-3-Opus** | **0.92 (0.89-0.94)** | **798 / 837 (95%, 94-97%)** | 362 / 1199 (30%, 28-33%) | **0.96 (0.93-0.98)** | **678 / 695 (98%, 96-99%)** | 350 / 1045 (33%, 31-36%) |
| **Claude-2** | **0.88 (0.85-0.91)** | **703 / 753 (93%, 92-95%)** | 479 / 1232 (39%, 36-42%) | **0.90 (0.87-0.93)** | **598 / 633 (94%, 93-96%)** | 434 / 1067 (41%, 38-44%) |
| GPT-4 | 0.83 (0.80-0.87) | 793 / 872 (91%, 89-93%) | 327 / 1199 (27%, 25-30%) | 0.81 (0.77-0.85) | 697 / 775 (90%, 88-92%) | 259 / 1034 (25%, 22-28%) |
| **GPT-3.5** | **0.93 (0.90-0.96)** | **563 / 587 (96%, 94-97%)** | 645 / 1232 (52%, 49-56%) | **0.90 (0.86-0.93)** | **503 / 535 (94%, 92-96%)** | 532 / 1067 (50%, 46-53%) |
| **Mixtral-8x22B** | **0.91 (0.87-0.94)** | **643 / 679 (95%, 93-96%)** | **542 / 1221 (44%, 41-48%)** | **0.93 (0.90-0.96)** | **535 / 556 (96%, 95-98%)** | **500 / 1056 (47%, 43-51%)** |

| PRECIS-2 (9 items × 56 publications = up to 504 ratings) |  |  |  |  |  |  |
| --- | --- | --- | --- | --- | --- | --- |
| *Human Rater 1 alone* | 0.57 (0.48-0.65) | 379 / 504 (75%, 70-80%) |  | 0.55 (0.45-0.65) | 369 / 504 (73%, 68-78%) |  |
| Claude-3-Opus | 0.73 (0.62-0.83) | 106 / 127 (83%, 77-90%) | 377 / 504 (75%, 72-78%) | 0.56 (0.41-0.70) | 102 / 137 (74%, 67-82%) | 367 / 504 (73%, 69-77%) |
| Claude-2 | 0.56 (0.35-0.76) | 101 / 126 (80%, 72-88%) | 378 / 504 (75%, 71-79%) | 0.56 (0.41-0.70) | 108 / 138 (78%, 71-85%) | 366 / 504 (73%, 68-77%) |
| GPT-4 | 0.63 (0.49-0.76) | 98 / 123 (80%, 72-87%) | 381 / 504 (76%, 72-80%) | 0.64 (0.50-0.78) | 101 / 128 (79%, 70-87%) | 376 / 504 (75%, 69-79%) |
| **GPT-3.5** | 0.68 (0.49-0.83) | 119 / 139 (86%, 79-91%) | 347 / 486 (71%, 68-75%) | 0.70 (0.56-0.83) | **124 / 143 (87%, 81-92%)** | 343 / 486 (71%, 66-75%) |
| Mixtral-8x22B | 0.61 (0.45-0.77) | 123 / 149 (83%, 76-89%) | 355 / 504 (70%, 66-75%) | 0.74 (0.63-0.84) | 126 / 147 (86%, 79-92%) | 357 / 504 (71%, 66-75%) |

**Supplementary Table 3:** Formatting and quoting accuracy.

| Rater | Publications unsuccessful | Successful retries | Minor formatting issues | Publications w/ quotes | Quotes | Perfect full text quotes | Perfect prompt quotes | Mean quote accuracy ± SD |
| --- | --- | --- | --- | --- | --- | --- | --- | --- |
| **PRISMA (n=112 publications)** |  |  |  |  |  |  |  |  |
| Claude-3-Opus | 3 / 112 (3%)<sup>a</sup> | 10 / 112 (9%)<sup>a</sup> | 3 / 109 (3%) | 108 / 109 (99%) | 1562 | 1140 / 1562 (73%) | 12 / 1562 (1%) | 98% ± 8% |
| Claude-2 | 0 / 112 (0%) | 0 / 112 (0%) | 6 / 112 (5%) | 79 / 112 (71%) | 557 | 359 / 557 (64%) | 51 / 557 (9%) | 95% ± 13% |
| GPT-4 | 3 / 112 (3%)<sup>b</sup> | 0 / 112 (0%) | 0 / 109 (0%) | 109 / 109 (100%) | 1829 | 1548 / 1829 (85%) | 3 / 1829 (0%) | 99% ± 4% |
| GPT-3.5 | 3 / 112 (3%)<sup>c</sup> | 7 / 112 (6%)<sup>c</sup> | 1 / 109 (1%) | 107 / 109 (98%) | 951 | 853 / 951 (90%) | 37 / 951 (4%) | 99% ± 4% |
| Mixtral-8x22B | 1 / 112 (1%)<sup>d</sup> | 14 / 112 (13%)<sup>d</sup> | 22 / 111 (20%) | 107 / 111 (96%) | 1616 | 880 / 1616 (54%) | 407 / 1616 (25%) | 97% ± 11% |
| **AMSTAR (n=112 publications)** |  |  |  |  |  |  |  |  |
| Claude-3-Opus | 3 / 112 (3%)<sup>a</sup> | 10 / 112 (9%)<sup>a</sup> | 3 / 109 (3%) | 109 / 109 (100%) | 861 | 621 / 861 (72%) | 26 / 861 (3%) | 97% ± 9% |
| Claude-2 | 0 / 112 (0%) | 0 / 112 (0%) | 6 / 112 (5%) | 110 / 112 (98%) | 480 | 326 / 480 (68%) | 64 / 480 (13%) | 97% ± 9% |
| GPT-4 | 3 / 112 (3%)<sup>b</sup> | 0 / 112 (0%) | 0 / 109 (0%) | 109 / 109 (100%) | 780 | 700 / 780 (90%) | 1 / 780 (0%) | 99% ± 4% |
| GPT-3.5 | 0 / 112 (0%) | 2 / 112 (2%)<sup>c</sup> | 49 / 112 (44%) | 112 / 112 (100%) | 847 | 809 / 847 (96%) | 7 / 847 (1%) | 99% ± 7% |
| Mixtral-8x22B | 1 / 112 (1%)<sup>d</sup> | 14 / 112 (13%)<sup>d</sup> | 22 / 111 (20%) | 108 / 111 (97%) | 830 | 643 / 830 (77%) | 44 / 830 (5%) | 98% ± 9% |
| **PRECIS-2 (n=56 publications)** |  |  |  |  |  |  |  |  |
| Claude-3-Opus | 0 / 56 (0%) | 0 / 56 (0%) | 0 / 56 (0%) | 56 / 56 (100%) | 533 | 306 / 533 (57%) | 0 / 533 (0%) | 95% ± 12% |
| Claude-2 | 0 / 56 (0%) | 0 / 56 (0%) | 2 / 56 (4%) | 56 / 56 (100%) | 479 | 355 / 479 (74%) | 56 / 479 (12%) | 99% ± 3% |
| GPT-4 | 0 / 56 (0%) | 0 / 56 (0%) | 0 / 56 (0%) | 56 / 56 (100%) | 562 | 480 / 562 (85%) | 1 / 562 (0%) | 99% ± 4% |
| GPT-3.5 | 2 / 56 (4%)<sup>c</sup> | 5 / 56 (9%)<sup>c</sup> | 23 / 54 (43%) | 54 / 54 (100%) | 566 | 533 / 566 (94%) | 0 / 566 (0%) | 99% ± 4% |
| Mixtral-8x22B | 0 / 56 (0%)<sup>d</sup> | 0 / 56 (0%)<sup>d</sup> | 0 / 56 (0%) | 56 / 56 (100%) | 564 | 463 / 564 (82%) | 0 / 564 (0%) | 99% ± 7% |

a: output blocked by content filtering policy, exceeded maximum output tokens, wrong format
b: publications too long for 32k context length when combining PRISMA and AMSTAR
c: stopped short, too verbose and thus exceeding context length, no response scores, nearly all NA
d: stopped short, nonsense gibberish, wrong format, empty, error

## Supplementary Prompt templates

### Supplementary prompt and example response for GPT-4 / PRECIS-2

#### System Prompt

You are an expert in clinical trial design and are tasked to assess the PRECIS-2 scores of a trial based on the full text of its publication. You have the briefing in the attached “Loudon 2015 shortened.txt” and “PRECIS Toolkit page 4.txt”. <Loudon 2015 shortened.txt><PRECIS Toolkit page 4.txt>

#### User Prompt

This is the full text to be assessed: <722.txt>

For each of the 9 domains of PRECIS-2, perform the following steps:

1. Extract 1-3 relevant quotes from the full text.
2. Explain your reasoning in 1 paragraph.
3. Give a score X from 1 (very explanatory) to 5 (very pragmatic) in square brackets like this: “Score: [X]”. If the full text does not contain enough information to assess a specific domain, report “Score: [NA]”.

Do not provide a final summary paragraph in the end.

#### Response (3/9 domains)

1. Eligibility criteria
  - Quote: “Participants had established CVD or an estimated five-year CVD risk of 15%, with indications for antiplatelet, statin and 2 blood pressure lowering drugs (‘combination treatment’). Participants randomized to the ‘polypill-based strategy’ received a polypill containing aspirin 75 mg, simvastatin 40 mg, lisinopril 10 mg and either atenolol 50 mg or hydrochlorothiazide 12.5 mg.”
  - Reasoning: The trial includes participants with established cardiovascular disease or a high risk of cardiovascular disease, which is a broad and inclusive group. However, the trial does exclude individuals who do not have indications for the combination treatment, which is a somewhat restrictive criterion.
  - Score: [4]
2. Recruitment
  - Quote: “A total of 623 participants were randomized from 33 centres (12 Aboriginal Medical Services) between January 2010 and May 2012.”
  - Reasoning: The trial recruited participants from a variety of centers, including Aboriginal Medical Services, which suggests a pragmatic approach to recruitment. However, the text does not provide detailed information about the recruitment process, such as whether participants were recruited through usual appointments or other methods.
  - Score: [NA]
3. Setting
  - Quote: “Participants attended the primary healthcare centres for trial assessments at randomization and 12 month intervals thereafter.”
  - Reasoning: The trial was conducted in primary healthcare centers, which is a typical setting for the delivery of the intervention in usual care. This suggests a pragmatic approach to the setting.
  - Score: [5]

### PRISMA and AMSTAR (combined for Claude-3-Opus, Claude-2, GPT-4, and Mixtral-8x22B)

#### System prompt

You are an expert in systematic reviews and are tasked to assess the methodological quality of a systematic review with the AMSTAR tool and its reporting quality with the PRISMA tool based on the full text of its publication. AMSTAR and PRISMA consist of the following items:

<AMSTAR>

A1. Was an ‘a priori’ design provided? The research question and inclusion criteria should be established before the conduct of the review. Note: Need to refer to a protocol, ethics approval, or pre-determined/a priori published research objectives to score a “yes.”

A2. Was there duplicate study selection and data extraction? There should be at least two independent data extractors and a consensus procedure for disagreements should be in place. Note: 2 people do study selection, 2 people do data extraction, consensus process or one person checks the other’s work.

A3. Was a comprehensive literature search performed? At least two electronic sources should be searched. The report must include years and databases used (e.g., Central, EMBASE, and MEDLINE). Key words and/or MESH terms must be stated and where feasible the search strategy should be provided. All searches should be supplemented by consulting current contents, reviews, textbooks, specialized registers, or experts in the particular field of study, and by reviewing the references in the studies found. Note: If at least 2 sources + one supplementary strategy used, select “yes” (Cochrane register/Central counts as 2 sources; a grey literature search counts as supplementary).

A4. Was the status of publication (i.e. grey literature) used as an inclusion criterion? The authors should state that they searched for reports regardless of their publication type. The authors should state whether or not they excluded any reports (from the systematic review), based on their publication status, language etc. Note: If review indicates that there was a search for “grey literature” or “unpublished literature,” indicate “yes.” SIGLE database, dissertations, conference proceedings, and trial registries are all considered grey for this purpose. If searching a source that contains both grey and non-grey, must specify that they were searching for grey/unpublished lit.

A5. Was a list of studies (included and excluded) provided? A list of included and excluded studies should be provided. Note: Acceptable if the excluded studies are referenced. If there is an electronic link to the list but the link is dead, select “no.”

A6. Were the characteristics of the included studies provided? In an aggregated form such as a table, data from the original studies should be provided on the participants, interventions and outcomes. The ranges of characteristics in all the studies analyzed e.g., age, race, sex, relevant socioeconomic data, disease status, duration, severity, or other diseases should be reported. Note: Acceptable if not in table format as long as they are described as above.

A7. Was the scientific quality of the included studies assessed and documented? ‘A priori’ methods of assessment should be provided (e.g., for effectiveness studies if the author(s) chose to include only randomized, double-blind, placebo controlled studies, or allocation concealment as inclusion criteria); for other types of studies alternative items will be relevant. Note: Can include use of a quality scoring tool or checklist, e.g., Jadad scale, risk of bias, sensitivity analysis, etc., or a description of quality items, with some kind of result for EACH study (“low” or “high” is fine, as long as it is clear which studies scored “low” and which scored “high”; a summary score/range for all studies is not acceptable).

A8. Was the scientific quality of the included studies used appropriately in formulating conclusions? The results of the methodological rigor and scientific quality should be considered in the analysis and the conclusions of the review, and explicitly stated in formulating recommendations. Note: Might say something such as “the results should be interpreted with caution due to poor quality of included studies.” Cannot score “yes” for this question if scored “no” for question A7.

A9. Were the methods used to combine the findings of studies appropriate? For the pooled results, a test should be done to ensure the studies were combinable, to assess their homogeneity (i.e., Chi-squared test for homogeneity, I2). If heterogeneity exists a random effects model should be used and/or the clinical appropriateness of combining should be taken into consideration (i.e., is it sensible to combine?). Note: Indicate “yes” if they mention or describe heterogeneity, i.e., if they explain that they cannot pool because of heterogeneity/variability between interventions.

A10. Was the likelihood of publication bias assessed? An assessment of publication bias should include a combination of graphical aids (e.g., funnel plot, other available tests) and/or statistical tests (e.g., Egger regression test, Hedges-Olken). Note: If no test values or funnel plot included, score “no”. Score “yes” if mentions that publication bias could not be assessed because there were fewer than 10 included studies.

A11. Was the conflict of interest included? Potential sources of support should be clearly acknowledged in both the systematic review and the included studies. Note: To get a “yes,” must indicate source of funding or support for the systematic review AND for each of the included studies..

</AMSTAR>

<PRISMA>

P1. Title: Identify the report as a systematic review, meta-analysis, or both.

P2. Abstract / Structured summary: Provide a structured summary including, as applicable: background; objectives; data sources; study eligibility criteria, participants, and interventions; study appraisal and synthesis methods; results; limitations; conclusions and implications of key findings; systematic review registration number.

P3. Introduction / Rationale: Describe the rationale for the review in the context of what is already known.

P4. Introduction / Objectives: Provide an explicit statement of questions being addressed with reference to participants, interventions, comparisons, outcomes, and study design (PICOS).

P5. Methods / Protocol and registration: Indicate if a review protocol exists, if and where it can be accessed (e.g., Web address), and, if available, provide registration information including registration number.

P6. Methods / Eligibility criteria: Specify study characteristics (e.g., PICOS, length of follow-up) and report characteristics (e.g., years considered, language, publication status) used as criteria for eligibility, giving rationale.

P7. Methods / Information sources: Describe all information sources (e.g., databases with dates of coverage, contact with study authors to identify additional studies) in the search and date last searched.

P8. Methods / Search: Present full electronic search strategy for at least one database, including any limits used, such that it could be repeated.

P9. Methods / Study selection: State the process for selecting studies (i.e., screening, eligibility, included in systematic review, and, if applicable, included in the meta-analysis).

P10. Methods / Data collection process: Describe method of data extraction from reports (e.g., piloted forms, independently, in duplicate) and any processes for obtaining and confirming data from investigators.

P11. Methods / Data items: List and define all variables for which data were sought (e.g., PICOS, funding sources) and any assumptions and simplifications made.

P12. Methods / Risk of bias in individual studies: Describe methods used for assessing risk of bias of individual studies (including specification of whether this was done at the study or outcome level), and how this information is to be used in any data synthesis.

P13. Methods / Summary measures: State the principal summary measures (e.g., risk ratio, difference in means).

P14. Methods / Synthesis of results: Describe the methods of handling data and combining results of studies, if done, including measures of consistency (e.g., I2) for each meta-analysis.

P15. Methods / Risk of bias across studies: Specify any assessment of risk of bias that may affect the cumulative evidence (e.g., publication bias, selective reporting within studies).

P16. Methods / Additional analyses: Describe methods of additional analyses (e.g., sensitivity or subgroup analyses, meta-regression), if done, indicating which were pre-specified.

P17. Results / Study selection: Give numbers of studies screened, assessed for eligibility, and included in the review, with reasons for exclusions at each stage, ideally with a flow diagram.

P18. Results / Study characteristics: For each study, present characteristics for which data were extracted (e.g., study size, PICOS, follow-up period) and provide the citations.

P19. Results / Risk of bias within studies: Present data on risk of bias of each study and, if available, any outcome level assessment (see item P12).

P20. Results / Results of individual studies: For all outcomes considered (benefits or harms), present, for each study: (a) simple summary data for each intervention group (b) effect estimates and confidence intervals, ideally with a forest plot.

P21. Results / Synthesis of results: Present results of each meta-analysis done, including confidence intervals and measures of consistency.

P22. Results / Risk of bias across studies: Present results of any assessment of risk of bias across studies (see Item P15).

P23. Results / Additional analysis: Give results of additional analyses, if done (e.g., sensitivity or subgroup analyses, meta-regression [see Item P16]).

P24. Discussion / Summary of evidence: Summarize the main findings including the strength of evidence for each main outcome; consider their relevance to key groups (e.g., healthcare providers, users, and policy makers).

P25. Discussion / Limitations: Discuss limitations at study and outcome level (e.g., risk of bias), and at review-level (e.g., incomplete retrieval of identified research, reporting bias).

P26. Discussion / Conclusions: Provide a general interpretation of the results in the context of other evidence, and implications for future research.

P27. Funding: Describe sources of funding for the systematic review and other support (e.g., supply of data); role of funders for the systematic review.

</PRISMA>

#### User prompt

This is the full text to be assessed:

<FULLTEXT>

%FULLTEXT%

</FULLTEXT>

For each of the 11 AMSTAR questions (A1 to A11) perform the following steps:

1. Extract 1-3 relevant quotes from the full text.
2. Explain your reasoning in 1 sentence.
3. Respond to the question with either “[Yes]” if adequate, “[No]” if inadequate, or “[NA]” if not applicable or not relevant to the text (for example, combining data in quantitative synthesis (A9) or assessing publication bias (A11) in the context of a systematic review without a meta-analysis).

For each of the PRISMA items (P1 to P27) perform the following steps:

1. Extract 1-3 relevant quotes from the full text.
2. Explain your reasoning in 1 sentence.
3. Respond with either “[Yes]” if the item was reported, “[No]” if not reported, or “[NA]” if not applicable (for example, items P14, P15, P16, P21, P22, P23 in the context of a systematic review without a meta-analysis).

Before stopping, make sure you’ve processed all 11 AMSTAR questions and 27 PRISMA items. Do not provide a final summary paragraph in the end.

### PRISMA (GPT-3.**5)**

#### System prompt

You are an expert in systematic reviews and are tasked to assess the reporting quality of a systematic review with the PRISMA 2009 tool based on the full text of its publication. PRISMA 2009 consist of the following items:

---

\# PRISMA 2009

P1. Title: Identify the report as a systematic review, meta-analysis, or both.

---

#### User prompt

This is the full text to be assessed:

---

%FULLTEXT%

---

For each of the PRISMA items (P1 to P27) perform the following steps. Do not repeat the items’ names / instructions.

1. Extract one relevant short quote from the full text.
2. Explain your reasoning in one short sentence.
3. Respond with either “[Yes]” if the item was reported, “[No]” if not reported, or “[NA]” if not applicable (for example, items P14, P15, P16, P21, P22, P23 in the context of a systematic review without a meta-analysis).

Be as concise as possible and avoid unnecessary repetitions. Before stopping, make sure you’ve processed all 27 PRISMA items. Do not provide a final summary paragraph in the end.

### AMSTAR (GPT-3.**5)**

#### System prompt

You are an expert in systematic reviews and are tasked to assess the methodological quality of a systematic review with the AMSTAR tool based on the full text of its publication. AMSTAR consists of the following items:

---

\# AMSTAR

A6. Were the characteristics of the included studies provided? In an aggregated form such as a table, data from the original studies should be provided on the participants, interventions and outcomes. The ranges of characteristics in all the studies analyzed e.g., age, race, sex, relevant socioeconomic data, disease status, duration, severity, or other diseases should be reported. Note: Acceptable if not in table format as long as they are described as above. Paul “Follow-up poorly reported in general”

---

#### User prompt

This is the full text to be assessed:

---

%FULLTEXT%

---

For each of the 11 AMSTAR questions (A1 to A11) perform the following steps:

Before stopping, make sure you’ve processed all 11 AMSTAR questions. Do not provide a final summary paragraph in the end.

### PRECIS-2 (Claude-3-Opus, GPT-3.**5, and Mixtral-8x22B)**

#### System prompt

You are an expert in clinical trial design and are tasked to assess the PRECIS-2 scores of a trial based on the full text of its publication. You have the following briefing:

---

\# PRECIS-2 Domains

* Eligibility: To what extent are the participants in the trial similar to those who would receive this intervention if it was part of usual care? For example, score 5 for very pragmatic criteria essentially identical to those in usual care; score 1 for a very explanatory approach with lots of exclusions (e.g. those who don’t comply, respond to treatment, or are not at high risk for primary outcome, are children or elderly), or uses many selection tests not used in usual care.
* Recruitment: How much extra effort is made to recruit participants over and above what that would be used in the usual care setting to engage with patients? For example, score 5 for very pragmatic recruitment through usual appointments or clinic; score 1 for a very explanatory approach with targeted invitation letters, advertising in newspapers, radio plus incentives and other routes that would not be used in usual care.
* Setting: How different is the setting of the trial and the usual care setting? For example, score 5 for a very pragmatic choice using identical settings to usual care; score 1, for a very explanatory approach with only a single centre, or only specialised trial or academic centres.
* Organisation: How different are the resources, provider expertise and the organisation of care delivery in the intervention arm of the trial and those available in usual care? For example, score 5 for a very pragmatic choice that uses identical organisation to usual care; score 1 for a very explanatory approach if the trial increases staff levels, gives additional training, require more than usual experience or certification and increase resources.
* Flexibility (delivery): How different is the flexibility in how the intervention is delivered and the flexibility likely in usual care? For example, score 5 for a very pragmatic choice with identical flexibility to usual care; score 1 for a very explanatory approach if there is a strict protocol, monitoring and measures to improve compliance, with specific advice on allowed co-interventions and complications.
* Flexibility (adherence): How different is the flexibility in how participants must adhere to the intervention and the flexibility likely in usual care? For example, score 5 for a very pragmatic choice involving no more than usual encouragement to adhere to the intervention; score 1 for a very explanatory approach that involves exclusion based on adherence, and measures to improve adherence if found wanting. In some trials eg surgical trials where patients are being operated on or Intensive Care Unit trials where patients are being given IV drug therapy, this domain is not applicable as there is no compliance issue after consent has been given, so this score should be left blank.
* Follow-up: How different is the intensity of measurement and follow-up of participants in the trial and the likely follow-up in usual care? For example, score 5 for a very pragmatic approach with no more than usual follow up; score 1 for a very explanatory approach with more frequent, longer visits, unscheduled visits triggered by primary outcome event or intervening event, and more extensive data collection.
* Primary outcome: To what extent is the trial’s primary outcome relevant to participants? For example, score 5 for a very pragmatic choice where the outcome is of obvious importance to participants; score 1 for a very explanatory approach using a surrogate, physiological outcome, central adjudication or use assessment expertise that is not available in usual care, or the outcome is measured at an earlier time than in usual care.
* Primary analysis: To what extent are all data included in the analysis of the primary outcome? For example, score 5 for a very pragmatic approach using intention to treat with all available data; score 1 for a very explanatory analysis that excludes ineligible post-randomisation participants, includes only completers or those following the treatment protocol.

---

#### User prompt

This is the full text to be assessed:

---

%FULLTEXT%

---

For each of the 9 domains of PRECIS-2, perform the following steps:

Do not provide a final summary paragraph in the end.

### PRECIS-2 (Claude-2 and GPT-4)

#### System prompt

You are an expert in clinical trial design and are tasked to assess the PRECIS-2 scores of a trial based on the full text of its publication. You have the briefing in the attached “Loudon 2015 [GPT-4: shortened].pdf” and “PRECIS Toolkit [GPT-4: page 4].pdf”

#### User prompt

This is the full text to be assessed: <ID.txt>

For each of the 9 domains of PRECIS-2, perform the following steps:

Do not provide a final summary paragraph in the end.

## Supplementary Figures

Supplementary Figures can be viewed online at:

https://timwoelfle.github.io/Evidence-Appraisal-AI/

